# The Association Between Dietary Changes to Lose Weight and Mental Health Status in the National Health and Nutrition Examination Survey, 2005-2006

**DOI:** 10.1101/2024.02.01.24302138

**Authors:** Jihyun Jane Min, Keeyoon Noh, Sojeong Nam, Alejandra Ellison-Barnes

## Abstract

**Background:** Weight loss is a common endeavor taken by millions of residents annually in the U.S.—a country with a 31% overweight and 42% obesity rate. Weight loss is associated with numerous physical health benefits, such as better cardiovascular health. However, unhealthy weight loss strategies can cause adverse mental health effects. Past research has focused on how weight loss strategies affect the mental health of a general population—rather than those who are trying to lose weight—or has investigated a diverse array of weight loss strategies. This paper explores how dietary changes for weight loss are associated with mental health, specifically through several variables used in the Patient Health Questionnaire-9 (PHQ9), which measures depression severity. We hypothesize that eating less and skipping meals will be associated with poorer mental health status, while eating fewer carbohydrates, eating less fat, and drinking more water will be associated with better mental health status.

**Methods and Findings:** This study uses the U.S. 2005-2006 National Health and Nutrition Examination Survey (NHANES) 2005. Univariate (descriptive statistics), bivariate (correlation coefficient), and multivariate (ordinal logistic regression) analyses were performed. The main results show that ‘skipped meals’ was positively associated with ‘feeling bad about yourself,’ ‘feeling down, depressed, or hopeless,’ and ‘little interest in doing things.’ ‘Ate fewer carbohydrates’ was negatively associated with the ‘little interest in doing things.’

**Conclusion:** Differing dietary changes used for weight loss, particularly skipping meals and eating fewer carbohydrates, are associated with differences in mental health status. Health care professionals providing weight loss guidance should be cognizant of patients’ baseline mental health and the potential for changes in mental health with different dietary strategies. Future research employing a longitudinal approach to determine whether there is evidence of a causal relationship between these and other dietary strategies and subsequent mental health outcomes.

## Introduction

With approximately 42% of its population being obese and 31% overweight, the United States holds millions of individuals attempting weight loss in a given year [1]. In fact, in 2013-2016, one-half of the U.S. population attempted to lose weight within the prior year [2]. Weight loss can reduce the likelihood of the long-term health consequences associated with obesity— including cancer, diabetes, and cardiovascular disease—and has added benefits, such as better physical mobility [1,3].

Due to its benefits, weight loss has been recommended by physicians at a rate of about 80% for individuals with obesity [4], but often advice of this nature has been vague and generic: simply suggesting that patients “eat less, do more” [5]. While this advice conveys the physiological core of weight loss, which is to have greater energy expenditure than energy intake from food consumption, without being given practical advice, individuals can respond to this recommendation through a variety of dietary changes that may impact not only their physical health but also their mental health. This advice also does not address the numerous determinants that factor into the success of weight loss, such as genetics, neighborhood, or food environment [6].

Prior literature demonstrates that weight loss and mental health have a multi-faceted relationship. A systematic review of several studies found nine that discovered an association between weight loss and improved mental health scores [7]. On the other hand, a different cohort study found that weight loss was linked to a greater likelihood of depression [8]. The leading author from the aforementioned cohort study posits that this is not due to the weight loss itself but the stresses of dieting, such as through the deprivation of food [9].

Several studies have examined how certain dietary changes that lead to weight loss are associated with changes in mental health. Many individuals attempting weight loss reduce carbohydrate intake since this macromolecule is found in energy-dense foods, such as pretzels or sugar-sweetened beverages, that contribute to weight gain [10]. A majority of studies conclude that excessive carbohydrate consumption is related to poorer mental health outcomes, particularly anxiety and depression [11–13]. However, a systematic review of controlled trials by Varaee et al. (2022) examining how low carbohydrate diets can influence depression and anxiety status concluded there was no significant association [14].

Fats are also often associated with weight gain, but studies examining their effects on mental health have mixed results. One experimental study found a linkage between increased susceptibility to depression and anxiety in mice that were fed high-fat diets, particularly with excess saturated fat, by impacting their neurological functions [15]. In contrast, low-fat diets were found to be related to greater anger and hostility for humans aged 20-37 [16]. There also seems to be a difference in results based on the type of fat consumed—while saturated fatty acids are positively associated with anxiety [17], unsaturated fatty acids, such as α-linolenic acid, were correlated with reduced likelihood for anxiety and depression [17,18]. Lastly, a few studies found no association between the level of fat consumption and mental health status or mood [19,20].

Drinking more water has also been shown to assist in weight loss through appetite suppression and reducing the average caloric intake for an individual [21]. Water can facilitate energy expenditure and lipolysis when the body breaks down fats for energy [22]. With its numerous physiological benefits, drinking water has been shown to greatly improve mood [23]. Conversely, dehydration was associated with increased tension/anxiety in studies examining this relationship for men (24).

Lastly, eating less and skipping meals allows for weight loss by reducing caloric consumption overall. In extreme forms, these methods pose risks for negative mental health outcomes as individuals are not consuming the micro and macronutrients essential for “preservation of normal brain function and [mental] well-being” [25, p.8]. A systematic review and meta-analysis found that breakfast skipping was related to depression, stress, and psychological distress (26). Additionally, for children, meal skipping was related to adolescent psychological complaints (27).

While abundant research has investigated how specific dietary changes can result in weight loss and impact people’s mental health, many have utilized general populations of people who may not be intentionally losing weight. For studies that focused on individuals with intentional weight loss, they do not focus on how specifically dietary changes are associated with mental health outcomes. Rather, they investigate a variety of changes that can lead to weight loss, such as lifestyle changes, weight-loss programs, or gastric bypass surgery (28–30). Noting this gap in the existing literature, we sought to determine whether a variety of dietary changes made by individuals seeking weight loss were associated with differing mental health status. Using a nationally representative sample, we evaluated the relationship between individuals’ dietary approaches to weight loss and their mental health status. We hypothesized that eating fewer carbohydrates, eating less fat, and drinking more water would be associated with better mental health status while eating less and skipping meals would be associated with poorer mental health status.

## Methods

### Data Source

This study used a nationally representative sample of the US population from the National Health and Nutrition Examination Survey (NHANES) 2005-2006, the results of which are publicly available. There is no special access to data from NHANES. Public use data files were used for the analysis and the dataset was downloaded directly from the CDC website. NHANES includes information on the health and nutrition status of adults and children, obtained through individual interviews and physical examinations (31). This survey was conducted by the National Center for Health Statistics (NCHS) periodically from 1971 to 1994. In 1999, the NHANES shifted to continuous data collection with a focus on a variety of health and nutrition measurements to meet emerging concerns [31]. The survey includes a nationally representative sample of approximately 5,000 participants each year across the US, with an oversampling of African Americans and Mexican Americans to provide accurate estimates for those populations [31]. We included adults aged 20 years and older and excluded those who responded ‘don’t know’ and/or ‘refuse’ for the variables of interest.

### Ethics Statement

All protocols were reviewed and approved by the NCHS Ethics Review Board with the U.S. Department of Health and Human Services Policy for Protection of Human Research Subjects before NHANES was implemented (32). Furthermore, this study was acknowledged by the Johns Hopkins University Institutional Review Board as Not Human Subjects Research (IRB00404102) and as a Quality Improvement project. Formal consent was not obtained as the dataset had all patient identifiers removed.

### Measures

*Mental health variables*. To measure overall mental health conditions, we selected four questions from the dataset. The first question was, “Over the last 2 weeks, how often have you been bothered by feeling bad about yourself?” Participants answered by selecting from ‘not at all (0),’ ‘several days (1),’ ‘more than half the days (2),’ and ‘nearly every day (3).’ The second question was, “Over the last 2 weeks, how often have you been bothered by feeling down, depressed, or hopeless?” Participants answered by selecting from ‘not at all (0),’ ‘several days (1),’ ‘more than half the days (2),’ and ‘nearly every day (3).’ The third question was, “Over the last 2 weeks, how often have you been bothered by the following problems: little interest or pleasure in doing things? Would you say…” Participants answered by selecting from ‘not at all (0),’ ‘several days (1),’ ‘more than half the days (2), and ‘nearly every day (3).’ The fourth question was, “how many days during the past 30 days has your mental health not been good?” To answer, participants directly provided the number of days. The first three survey questions are part of the Patient Health Questionnaire-9 (PHQ9) that measures depression severity [32].

*Dietary variables.* Participants were asked, “How did you try to lose weight?” and were provided with a card with options that included, ‘ate less fat,’ ‘ate fewer carbohydrates,’ ‘drank a lot of water,’ ‘ate less food,’ and ‘skipped meals,’ among other choices. Respondents were prompted to select all that applied.

*Covariates.* We controlled for participants’ age, sex, race, educational level, and annual household income.

*A summary of the variables and the questions asked for this study can be found in the tables below:*

| Description | Question asked |
| --- | --- |
| No. of days mental health was not good | Now thinking about {your/SP's} mental health, which includes stress, depression, and problems with emotions, for how many days during the past 30 days was {your/his/her} mental health not good? |
| Feeling bad about yourself | Over the last 2 weeks, how often have you been bothered by the following problems:] feeling bad about yourself - or that you are a failure or have let yourself or your family down? |
| Feeling down, depressed, or hopeless | Over the last 2 weeks, how often have you been bothered by the following problems: feeling down, depressed, or hopeless? |
| Little interest in doing things | Over the last 2 weeks, how often have you been bothered by the following problems: little interest or pleasure in doing things? Would you say... |

### Statistical Analysis

For the study, the quantitative software IBM SPSS was used for univariate (descriptive statistics), bivariate (correlation coefficient), and multivariate (ordinal logistic regression and negative binomial regression models) analyses.

## Results

We included a total of 3,975 participants in our sample (Table 1). The average age was 47.54 years old (SD = 17.85), ranging from 20 to 84.92 years; 48.3% of participants were male (n = 1,919), and 51.7% were female (n = 2,056). In terms of race/ethnicity, 50.9% of participants were non-Hispanic White (n = 2,022), 22.6% were non-Hispanic Black (n = 898), 22.7% were Mexican American (n = 904), and 3.8% identified as other race or multi/bi-racial (n = 151). Education levels varied among our sample: 25.9% had less than a high school education (n = 1,030), 23.9% had a high school diploma or GED (n = 952), 29.1% had some college or an AA degree (n = 1,158), and 21% had a college degree or higher (n = 835). The average annual household income was $38,199.68 (SD = $12,349.53).

**Table 1.** Descriptive statistics for the variables in the analyses (n = 3,975; NHANES 2005-2006).

| Variables | n | % | Mean | SD | Min | Max |
| --- | --- | --- | --- | --- | --- | --- |
| **Dependent Variables** |  |  |  |  |  |  |
| Feeling bad about yourself |  |  |  |  |  |  |
| Not at all | 3,409 | 85.8 |  |  |  |  |
| Several days | 407 | 10.2 |  |  |  |  |
| More than half the days | 83 | 2.1 |  |  |  |  |
| Nearly every day | 76 | 1.9 |  |  |  |  |
| Feeling down, depressed, or hopeless |  |  |  |  |  |  |
| Not at all | 3,111 | 78.3 |  |  |  |  |
| Several days | 618 | 15.5 |  |  |  |  |
| More than half the days | 152 | 3.8 |  |  |  |  |
| Nearly every day | 94 | 2.4 |  |  |  |  |
| Little interest in doing things |  |  |  |  |  |  |
| Not at all | 3,188 | 80.2 |  |  |  |  |
| Several days | 534 | 13.4 |  |  |  |  |
| More than half the days | 153 | 3.8 |  |  |  |  |
| Nearly every day | 100 | 2.5 |  |  |  |  |
| N. of days mental health was not good | 3975 | 100 | 3.56 | 7.58 | 0 | 30 |
| **Independent Variables** |  |  |  |  |  |  |
| Healthy Diet |  |  |  |  |  |  |
| Drank much water (yes) | 831 | 20.9 |  |  |  |  |
| Ate less fat to lose weight (yes) | 765 | 19.2 |  |  |  |  |
| Ate few carbs (yes) | 561 | 14.1 |  |  |  |  |
| Unhealthy Diet |  |  |  |  |  |  |
| Ate less food to lose weight (yes) | 1,179 | 29.7 |  |  |  |  |
| Skipped meals (yes) | 370 | 9.3 |  |  |  |  |
| Covariates |  |  |  |  |  |  |
| Age |  |  | 47.54 | 17.85 | 20 | 84.92 |
| Sex |  |  |  |  |  |  |
| Male | 1,919 | 48.3 |  |  |  |  |
| Female | 2,056 | 51.7 |  |  |  |  |
| Race |  |  |  |  |  |  |
| White | 2,022 | 50.9 |  |  |  |  |
| African American | 898 | 22.6 |  |  |  |  |
| Hispanic | 904 | 22.7 |  |  |  |  |
| Other race | 151 | 3.8 |  |  |  |  |
| Educational level |  |  |  |  |  |  |
| Less than high school | 1,030 | 25.9 |  |  |  |  |
| High school/GED | 952 | 23.9 |  |  |  |  |
| College level/AA | 1,158 | 29.1 |  |  |  |  |
| College or above | 835 | 21 |  |  |  |  |
| Annual household income | | | \$38,199.68 | \$12,349.53 | | |

For the mental health variables, about 14% of the participants reported that they ‘[felt] bad about [themselves]’ for several days or longer over the last two weeks. 22% reported ‘feeling down, depressed, or hopeless’ for several days or longer, and about 20% had ‘little interest in doing things’ for several days or longer. The mean ‘number of days mental health was not good’ during the past month was 3.56.

In terms of dietary changes to lose weight, the number of respondents who marked ‘yes’ for the options ‘drank a lot of water,’ ‘ate less fat’ and ‘ate fewer carbohydrates’ were 20.9% (n = 831), 19.2% (n = 765), and 14.1% (n = 561), respectively. For the other dietary variables, the numbers of respondents who responded ‘yes’ to ‘ate less food’ and ‘skipped meals’ were 29.7% (n = 1,179) and 9.3% (n = 370), respectively.

In the bivariate analysis (Table 2), all of the dietary changes were significantly correlated with at least one mental health status. ‘Ate less food’ was significantly correlated with ‘number of days mental health was not good.’ ‘Ate fewer carbohydrates’ and ‘ate less fat’ were significantly associated with ‘little interest in doing things.’ ‘Drank a lot of water’ was associated with ‘feeling down, depressed, or hopeless,’ and ‘number of days mental health was not good.’ *‘*Skipped meals*’* was associated with ‘feeling bad about yourself,’ ‘feeling down, depressed, or hopeless,’ ‘little interest in doing things,’ and ‘number of days mental health was not goo

**Table 2.** Bivariates analysis among the variables.

|  | (a) | (b) | (c) | (d) | (e) | (f) | (g) | (h) | (i) | (j) | (k) | (l) | (m) | (n) |
| --- | --- | --- | --- | --- | --- | --- | --- | --- | --- | --- | --- | --- | --- | --- |
| (a)Feeling bad about yourself | 1 |  |  |  |  |  |  |  |  |  |  |  |  |  |
| (b)Feeling down, depressed, or hopeless | .583** | 1 |  |  |  |  |  |  |  |  |  |  |  |  |
| (c)Little interest in doing things | .414** | .515** | 1 |  |  |  |  |  |  |  |  |  |  |  |
| (d)No. of days mental health was not good | .476** | .599** | .398** | 1 |  |  |  |  |  |  |  |  |  |  |
| (e)Drank much water | .016 | .029* | .010 | .057** | 1 |  |  |  |  |  |  |  |  |  |
| (f)Ate less fat | .009 | -.004 | -.009 | .031* | .486** | 1 |  |  |  |  |  |  |  |  |
| (g)Ate fewer carbs | -.003 | -.019 | -.042** | .021 | .420** | .457** | 1 |  |  |  |  |  |  |  |
| (h)Ate less food to lose weight | .030* | .016 | .015 | .056** | .528** | .493** | .388** | 1 |  |  |  |  |  |  |
| (i)Skipped meals | .053** | .052** | .055** | .069** | .369** | .264** | .196** | .388** | 1 |  |  |  |  |  |
| (j)Age | -.020 | -.023 | -.001 | -.051** | -.077** | .001 | -.016 | .005 | -.059** | 1 |  |  |  |  |
| (k)Gender | .062** | .077** | .035* | .110** | .139** | .085** | .088** | .122** | .071** | -.081** | 1 |  |  |  |
| (l)Race & ethnicity | -.006 | -.019 | .012 | .037** | .036** | .020* | .033** | .028** | .049** | .077** | -.001 | 1 |  |  |
| (m)Education level | -.070** | -.111** | -.111** | -.043** | .147** | .111** | .150** | .096** | .030* | -.095** | .047** | .262** | 1 |  |
| (n)Household income | -.132** | -.159** | -.115** | -.080** | .065** | .067** | .107** | .046** | -.002 | -.127** | -.044** | .076** | .415** | 1 |

*Note. \* $p < 0.05$ , \*\* $p < 0$ .*

Ordinal logistic regression estimates predicting the three mental health variables measured by a Likert scale: ‘feeling bad about yourself,’ ‘feeling down, depressed, or hopeless,’ and ‘little interest in doing things’ are shown in Table 3.

**Table 3.**
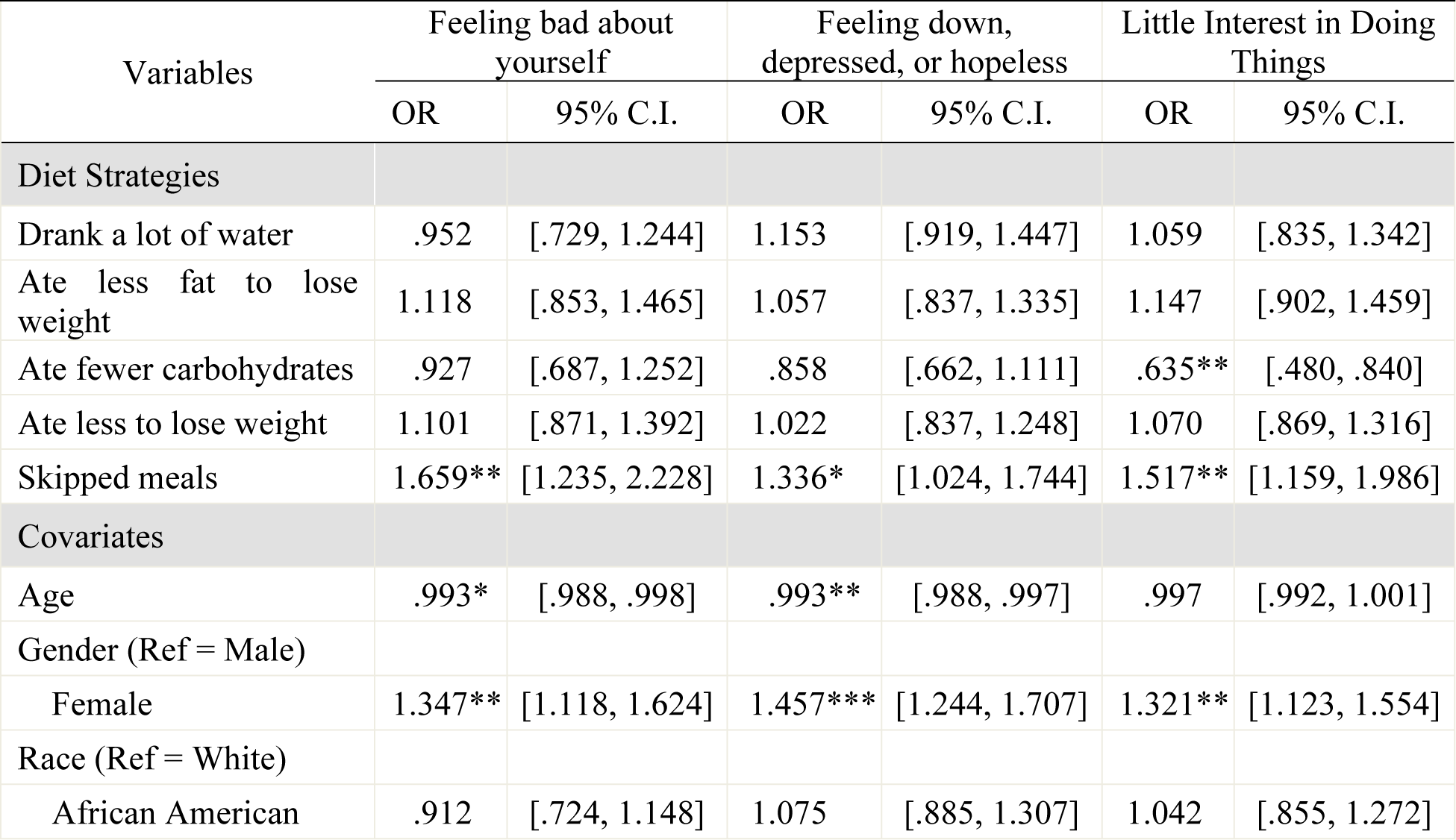
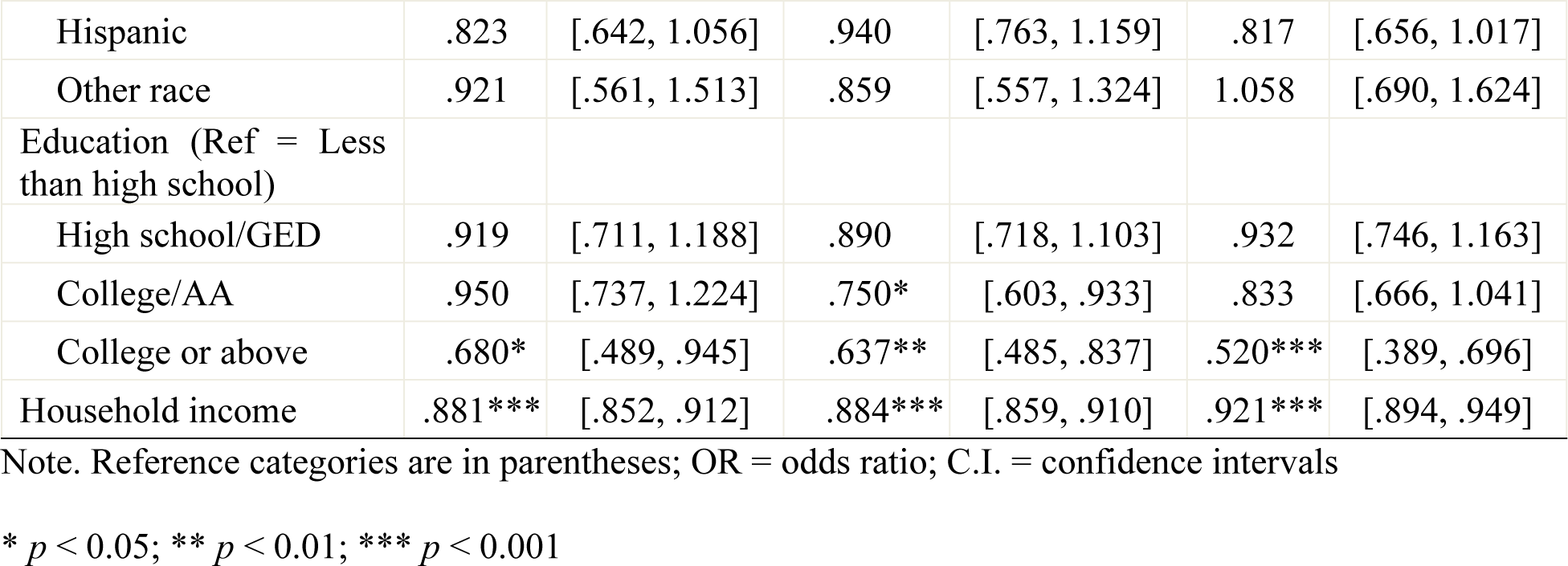
Associations between Feeling Bad about Yourself, Feeling Down, Depressed, or Hopeless, and Little Interest in Doing Things and diet strategies as assessed from NHANES 2005-2006.

| Variables | Feeling bad about yourself |  | Feeling down, depressed, or hopeless |  | Little Interest in Doing Things |  |
| --- | --- | --- | --- | --- | --- | --- |
|  | OR | 95% C.I. | OR | 95% C.I. | OR | 95% C.I. |
| **Diet Strategies** |  |  |  |  |  |  |
| Drank a lot of water | .952 | [.729, 1.244] | 1.153 | [.919, 1.447] | 1.059 | [.835, 1.342] |
| Ate less fat to lose weight | 1.118 | [.853, 1.465] | 1.057 | [.837, 1.335] | 1.147 | [.902, 1.459] |
| Ate fewer carbohydrates | .927 | [.687, 1.252] | .858 | [.662, 1.111] | .635** | [.480, .840] |
| Ate less to lose weight | 1.101 | [.871, 1.392] | 1.022 | [.837, 1.248] | 1.070 | [.869, 1.316] |
| Skipped meals | 1.659** | [1.235, 2.228] | 1.336* | [1.024, 1.744] | 1.517** | [1.159, 1.986] |
| **Covariates** |  |  |  |  |  |  |
| Age | .993* | [.988, .998] | .993** | [.988, .997] | .997 | [.992, 1.001] |
| Gender (Ref = Male) |  |  |  |  |  |  |
| Female | 1.347** | [1.118, 1.624] | 1.457*** | [1.244, 1.707] | 1.321** | [1.123, 1.554] |
| Race (Ref = White) |  |  |  |  |  |  |
| African American | .912 | [.724, 1.148] | 1.075 | [.885, 1.307] | 1.042 | [.855, 1.272] |
| Hispanic | .823 | [.642, 1.056] | .940 | [.763, 1.159] | .817 | [.656, 1.017] |
| Other race | .921 | [.561, 1.513] | .859 | [.557, 1.324] | 1.058 | [.690, 1.624] |
| Education (Ref = Less than high school) |  |  |  |  |  |  |
| High school/GED | .919 | [.711, 1.188] | .890 | [.718, 1.103] | .932 | [.746, 1.163] |
| College/AA | .950 | [.737, 1.224] | .750* | [.603, .933] | .833 | [.666, 1.041] |
| College or above | .680* | [.489, .945] | .637** | [.485, .837] | .520*** | [.389, .696] |
| Household income | .881*** | [.852, .912] | .884*** | [.859, .910] | .921*** | [.894, .949] |

Note. Reference categories are in parentheses; OR = odds ratio; C.I. = confidence intervals
\* $p < 0.05$ ; \*\* $p < 0.01$ ; \*\*\* $p < 0.001$

Participants who reported ‘skipped meals’ were more likely to endorse ‘feeling bad about yourself’ (OR = 1.659, p < .01). ‘Skipped meals’ was also positively associated with ‘feeling down, depressed, or hopeless’ (OR = 1.336, *p* < .05). Lastly, ‘skipped meals’ had a positive association with ‘little interest in doing things’ (OR = 1.517, *p* < .01).

‘Ate fewer carbohydrates had a positive association with ‘little interest in doing things’ (OR = .635, p < .01). In other words, individuals who reported that they ate fewer carbohydrates to lose weight were more likely to have little interest in doing things.

The results from a negative binomial regression model predicting ‘the number of days mental health was not good’ are shown in Table 4. None of the dietary variables was significantly associated with an increased odd of reporting more days of poor mental health.

## Discussion

In the analysis of the NHANES 2005-2006 dataset, we found varying associations between dietary changes and mental health status. In regression analysis controlling for age, sex, race, educational level, and annual household income, one of the variables, ‘skipped meals,’ was positively associated with ‘feeling bad about yourself,’ ‘feeling down, depressed, or hopeless,’ and ‘little interest in doing things.’ ‘Ate fewer carbohydrates’ was positively associated with ‘little interest in doing things.’ While the results support our hypothesis that skipping meals would have a negative association with mental health, they disagree with our initial claim that reduced carbohydrate intake would be correlated to positive mental health. The results also differ from our initial belief in that eating less fat, eating less, and drinking more water would show a significant association with the mental health variables. Our findings indicate that specific dietary changes to lose weight may be associated with differing mental health statuses, which may be relevant for healthcare professionals advising on weight loss strategies.

**Table 4.**
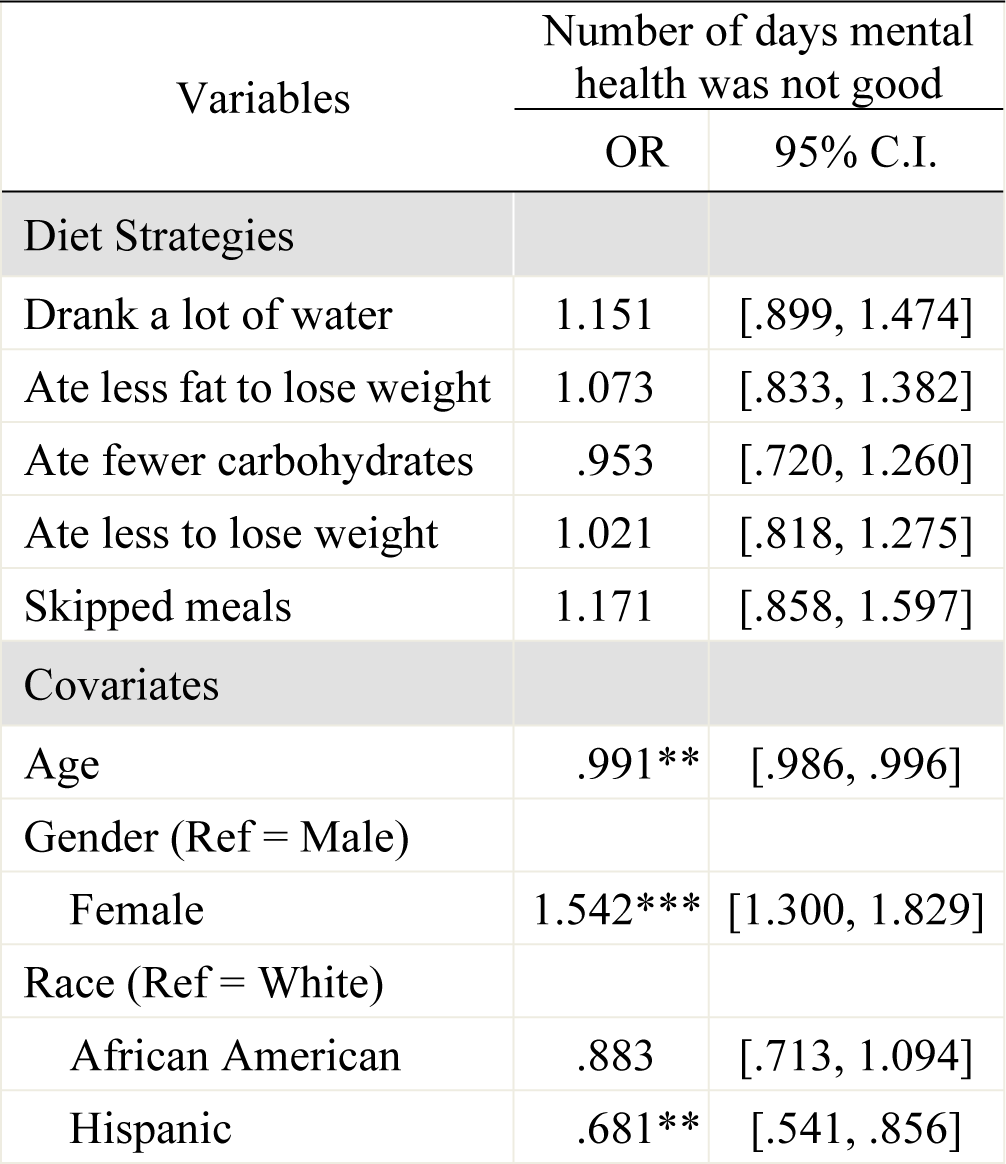
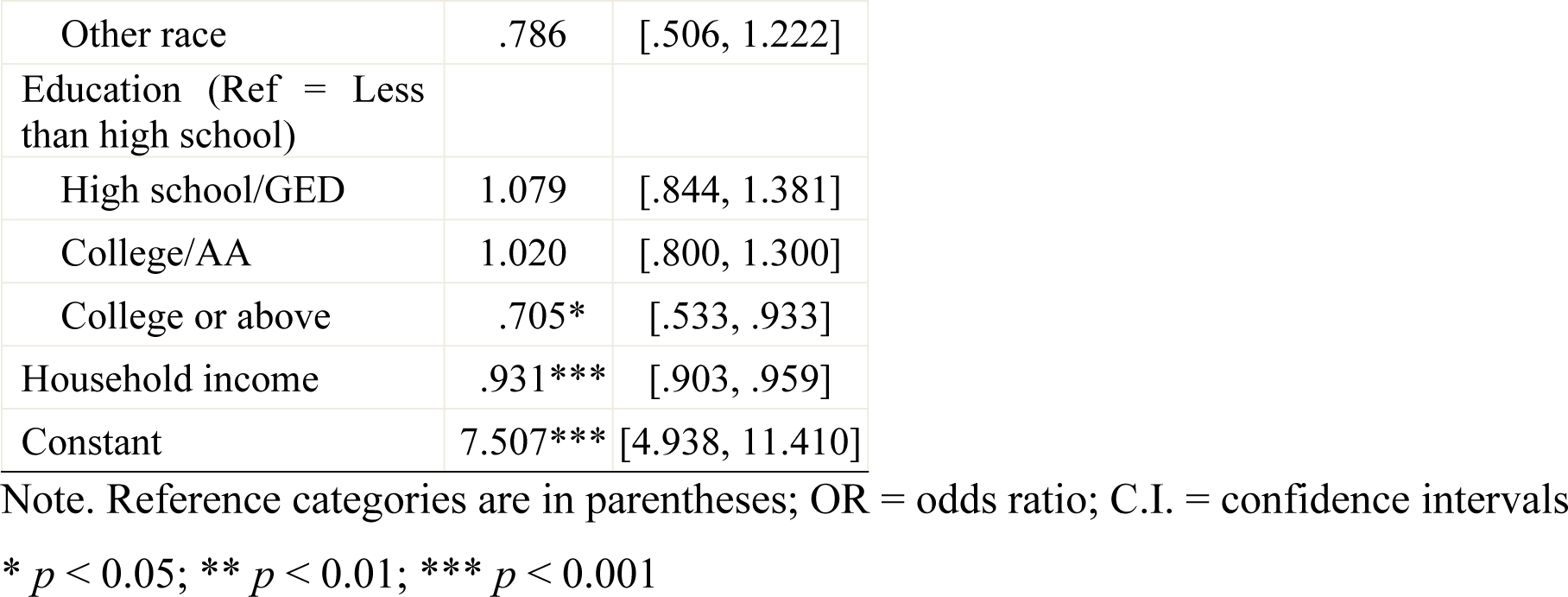
Associations between the Number of Days Mental Health was Not Good and diet strategies as assessed from NHANES 2005-2006.

Our results have several significant clinical implications. Our finding on the potential association between reduced carbohydrate intake and ‘little interest in doing things’ may be supported by prior literature that carbohydrates are essential in maintaining proper mental health [33]. Professionals should recommend that patients limit refined carbohydrates but maintain appropriate carbohydrate intake overall while proceeding with weight loss. Meanwhile, we found that ‘skipping meals’ is associated with poorer mental health status, which is consistent with prior studies’ results [26,34,35]. Skipping meals is known to be associated wth negative physical health consequences, including a reduced metabolism that can even promote weight gain when an individual consumes their typical food portions [36]. Thus, clinicians can emphasize how weight loss can be achieved through eating a healthy, balanced diet without skipping meals.

Our findings also indicate the necessity of integrating considerations for mental health in weight loss. To tailor weight loss strategies and interventions appropriately, it is crucial to incorporate an understanding of patients’ mental health. Clinicians should be cognizant of the diverse weight loss programs and interventions available, particularly those explicitly designed with mental health considerations. For instance, implementing interventions like a stress management program, as demonstrated by effectiveness in not only reducing weight or BMI but also decreasing depression and anxiety scores [37], can ensure that patients achieve their weight loss goals while safeguarding their mental health.

This study included regression analysis to examine the relationship between weight loss strategies and mental health status. While this analysis treated mental health status as a dependent variable, previous research employing regression analysis has also established a significant association between improvements in mental health symptoms during weight loss treatment and actual weight loss. This suggests that there may be meaningful implications in both directions between dietary changes and mental health [38,39]. Recognizing the association between certain strategies for weight loss and mental health status during the weight loss process underscores the imperative to discuss patients’ past and present mental health experiences. Identifying vulnerabilities or predispositions to adverse mental health symptoms allows clinicians to proactively recognize patients at risk during their weight loss journey. Incorporating additional assessments ensures that these individuals receive appropriate, mental health-informed care throughout the weight loss process.

## Limitations

There are several limitations of this study. The NHANES dataset is cross-sectional and observational, meaning we cannot define the directionality of the relationship between participants’ dietary changes and their mental health status and whether there is causality. Therefore, future studies should include longitudinal data and consider including potential mediators such as stress in their research designs. Next, while the 2005-2006 dataset contained questions that asked about people’s dietary changes in losing weight, more recent datasets did not include these questions, preventing this study from utilizing a more recent dataset that could be more generalizable to the current U.S. population. Even though NHANES is representative of the U.S. population, the 2005-2006 dataset also neglected the unique outcomes of Asian Americans or Native Americans by combining them under “other races or multi/bi-racial.” Future research exploring similar questions should incorporate more diverse racial and ethnic populations and observe their associations over time to yield results applicable to more diverse groups of people. Third, we could not differentiate the type of fat or carbohydrates the participants tried to reduce in their dietary changes. Prior studies have found that consuming more saturated fatty acids was associated with poorer mental health, while consuming more unsaturated fatty acids was associated with better mental health [17,18]; we could not examine the associations of different types of fats with mental health status because this information was not available in the dataset. Finally, our mental health variables captured mainly depression symptoms rather than the broad spectrum of mental illness. Future studies should utilize variables capturing other aspects of mental health.

Finally, measuring a mental construct with selective questions is problematic. It may not capture the construct sufficiently and accurately. For instance, as opposed to our last question, “How many days during the past 30 days has your mental health not been good?” the first three questions were more likely to show depressive symptoms or emotions, indicating different constructs (e.g., stress, depression, and problems with emotions). Therefore, future studies should design questions with high construct validity.

## Conclusion

Our results make clear that individuals seeking weight loss should receive support from healthcare professionals in determining the weight loss strategies that would be most appropriate while supporting maintenance of mental health. With practical advice, individuals can choose weight loss strategies that may be associated with improvements in both physical and mental health.

## Data Availability

https://wwwn.cdc.gov/nchs/nhanes/continuousnhanes/default.aspx?BeginYear=2005

## Acknowledgements

We would like to acknowledge Johns Hopkins University in their support for the publication funding for this paper. We also would like to acknowledge Dr. Keeyoon Noh’s time analyzing and interpreting the NHANES 2005-2006 dataset for our study.

## References

1. Centers for Disease Control and Prevention. Health Effects of Overweight and Obesity [Internet]. 2022 [cited 2023 Sep 1]. Available from: https://www.cdc.gov/healthyweight/effects/index.html

2. Martin CB, Herrick KA, Sarafrazi N, Ogden CL. Attempts to Lose Weight Among Adults in the United States, 2013-2016. NCHS Data Brief. 2018 Jul;(313):1–8.

3. Safaei M, Sundararajan EA, Driss M, Boulila W, Shapi’i A. A systematic literature review on obesity: Understanding the causes & consequences of obesity and reviewing various machine learning approaches used to predict obesity. Comput Biol Med. 2021 Sep;136:104754.

4. Collins, K. Doctors on fat: Don’t ask, don’t tell [Internet]. nbcnews; 2003. Available from: https://www.nbcnews.com/id/wbna3679207

5. Tremblett M, Poon AYX, Aveyard P, Albury C. What advice do general practitioners give to people living with obesity to lose weight? A qualitative content analysis of recorded interactions. Fam Pract. 2022 Dec 13;cmac137.

6. Lee A, Cardel M, Donahoo WT. Social and Environmental Factors Influencing Obesity. In: Feingold KR, Anawalt B, Blackman MR, Boyce A, Chrousos G, Corpas E, et al., editors. Endotext [Internet]. South Dartmouth (MA): MDText.com, Inc.; 2000 [cited 2023 Dec 27]. Available from: http://www.ncbi.nlm.nih.gov/books/NBK278977/

7. Lasikiewicz N, Myrissa K, Hoyland A, Lawton CL. Psychological benefits of weight loss following behavioural and/or dietary weight loss interventions. A systematic research review. Appetite. 2014 Jan;72:123–37.

8. Jackson SE, Steptoe A, Beeken RJ, Kivimaki M, Wardle J. Psychological Changes following Weight Loss in Overweight and Obese Adults: A Prospective Cohort Study. Franken IHa, editor. PLoS ONE. 2014 Aug 6;9(8):e104552.

9. Berenson T. Losing Weight Could Make You Depressed, Study Says [Internet]. TIME; 2014. Available from: https://time.com/3092086/weight-loss-depression/

10. Dam RM van, Seidell JC. Carbohydrate intake and obesity. Eur J Clin Nutr. 2007;61:S75–99.

11. Christensen L. The effect of carbohydrates on affect. Nutr Burbank Los Angel Cty Calif. 1997 Jun;13(6):503–14.

12. Gopinath B, Flood VM, Burlutksy G, Louie JCY, Mitchell P. Association between carbohydrate nutrition and prevalence of depressive symptoms in older adults. Br J Nutr. 2016;116(12):2109–14.

13. Kose J, Duquenne P, Robert M, Debras C, Galan P, Péneau S, et al. Associations of overall and specific carbohydrate intake with anxiety status evolution in the prospective NutriNet-Santé population-based cohort. Sci Rep. 2022 Dec 14;12(1):21647.

14. Varaee H, Darand M, Hassanizadeh S, Hosseinzadeh M. Effect of low-carbohydrate diet on depression and anxiety: A systematic review and meta-analysis of controlled trials. J Affect Disord. 2023 Mar;325:206–14.

15. Zemdegs J, Quesseveur G, Jarriault D, Pénicaud L, Fioramonti X, Guiard BP. High-fat diet-induced metabolic disorders impairs 5-HT function and anxiety-like behavior in mice. Br J Pharmacol. 2016 Jul;173(13):2095–110.

16. Wells AS, Read NW, Laugharne JDE, Ahluwalia NS. Alterations in mood after changing to a low-fat diet. Br J Nutr. 1998 Jan;79(1):23–30.

17. Fatemi F, Siassi F, Qorbani M, Sotoudeh G. Higher dietary fat quality is associated with lower anxiety score in women: a cross-sectional study. Ann Gen Psychiatry. 2020 Dec;19(1):14.

18. Daley C, Patterson A, Sibbritt D, MacDonald-Wicks L. Unsaturated fat intakes and mental health outcomes in young women from the Australian Longitudinal Study on Women’s Heath. Public Health Nutr. 2015 Feb;18(3):546–53.

19. Wardle J, Rogers P, Judd P, Taylor MA, Rapoport L, Green M, et al. Randomized trial of the effects of cholesterol-lowering dietary treatment on psychological function. Am J Med. 2000 May;108(7):547–53.

20. Wilson JJ, McMullan I, Blackburn NE, Klempel N, Yakkundi A, Armstrong NC, et al. Changes in dietary fat intake and associations with mental health in a UK public sample during the COVID-19 pandemic. J Public Health Oxf Engl. 2021 Dec 10;43(4):687–94.

21. Vij VAK, Joshi AS. Effect of excessive water intake on body weight, body mass index, body fat, and appetite of overweight female participants. J Nat Sci Biol Med. 2014 Jul;5(2):340–4.

22. Boschmann M, Steiniger J, Hille U, Tank J, Adams F, Sharma AM, et al. Water-induced thermogenesis. J Clin Endocrinol Metab. 2003 Dec;88(12):6015–9.

23. Pross N, Demazières A, Girard N, Barnouin R, Metzger D, Klein A, et al. Effects of changes in water intake on mood of high and low drinkers. PloS One. 2014;9(4):e94754.

24. Ganio MS, Armstrong LE, Casa DJ, McDermott BP, Lee EC, Yamamoto LM, et al. Mild dehydration impairs cognitive performance and mood of men. Br J Nutr. 2011 Nov;106(10):1535–43.

25. Muscaritoli M. The Impact of Nutrients on Mental Health and Well-Being: Insights From the Literature. Front Nutr. 2021;8:656290.

26. Zahedi H, Djalalinia S, Sadeghi O, Zare Garizi F, Asayesh H, Payab M, et al. Breakfast consumption and mental health: a systematic review and meta-analysis of observational studies. Nutr Neurosci. 2022 Jun;25(6):1250–64.

27. Azemati B, Heshmat R, Qorbani M, Ahadi Z, Azemati A, Shafiee G, et al. Association of meal skipping with subjective health complaints in children and adolescents: the CASPIAN-V study. Eat Weight Disord EWD. 2020 Feb;25(1):241–6.

28. Blaine BE, Rodman J, Newman JM. Weight Loss Treatment and Psychological Well-being: A Review and Meta-analysis. J Health Psychol. 2007 Jan;12(1):66–82.

29. Chaitoff A, Swetlik C, Ituarte C, Pfoh E, Lee LL, Heinberg LJ, et al. Associations Between Unhealthy Weight-Loss Strategies and Depressive Symptoms. Am J Prev Med. 2019 Feb;56(2):241–50.

30. Fabricatore AN, Wadden TA, Higginbotham AJ, Faulconbridge LF, Nguyen AM, Heymsfield SB, et al. Intentional weight loss and changes in symptoms of depression: a systematic review and meta-analysis. Int J Obes. 2011;35(11):1363–76.

31. ICPSR. National Health and Nutrition Examination Survey (NHANES), 2005-2006 (ICPSR 25504) [Internet]. Institute for Social Research University of Michigan; 2023. Available from: https://www.icpsr.umich.edu/web/ICPSR/studies/25504

32. U.S. Department of Health and Human Services Center for Disease Control and Prevention National Center for Health Statistics. National Health and Nutrition Examination Survey: Sample Design, 1999-2006. Available from https://wwwn.cdc.gov/nchs/nhanes/analyticguidelines.aspx

33. Kroenke K, Spitzer RL, Williams JB. The PHQ-9: validity of a brief depression severity measure. J Gen Intern Med. 2001 Sep;16(9):606–13.

34. Clemente-Suárez VJ, Mielgo-Ayuso J, Martín-Rodríguez A, Ramos-Campo DJ, Redondo-Flórez L, Tornero-Aguilera JF. The Burden of Carbohydrates in Health and Disease. Nutrients. 2022 Sep 15;14(18):3809.

35. Lee G, Han K, Kim H. Risk of mental health problems in adolescents skipping meals: The Korean National Health and Nutrition Examination Survey 2010 to 2012. Nurs Outlook. 2017;65(4):411–9.

36. Stanton R, Best T, Williams S, Vandelanotte C, Irwin C, Heidke P, et al. Associations between health behaviors and mental health in Australian nursing students. Nurse Educ Pract. 2021 May;53:103084.

37. Kuppersmith N, Kennedy C. Perils of Skipping Meals [Internet]. University of Louisville Research Foundation Inc.; 2005. Available from: https://louisville.edu/medicine/departments/familymedicine/files/L081611.pdf

38. Xenaki N, Bacopoulou F, Kokkinos A, Nicolaides NC, Chrousos GP, Darviri C. Impact of a stress management program on weight loss, mental health and lifestyle in adults with obesity: a randomized controlled trial. J Mol Biochem. 2018;7(2):78–84.

39. Alhalel N, Schueller SM, O’Brien MJ. Association of changes in mental health with weight loss during intensive lifestyle intervention: does the timing matter? Obes Sci Pract. 2018 Apr;4(2):153–8.

40. Payne ME, Porter Starr KN, Orenduff M, Mulder HS, McDonald SR, Spira AP, et al. Quality of Life and Mental Health in Older Adults with Obesity and Frailty: Associations with a Weight Loss Intervention. J Nutr Health Aging. 2018;22(10):1259– 65.

